# Soluble Glycoprotein 120 is associated with Coronary Artery Inflammation Measured by Pericoronary Fat Attenuation Index in People Living with HIV

**DOI:** 10.64898/2026.05.05.26352462

**Authors:** Pêngd-Wendé Habib Boussé Traoré, Kevin Boczar, Mehdi Benlarbi, Victoria Devine-Ducharme, Valerie Shirobokov, Bethlehem Mengesha, Jonathan Richard, Nicolas Chomont, Marc Messier-Peet, Annie Chamberland, Branka Vulesevic, Mohamed El-Far, Cécile Tremblay, Carl Chartrand-Lefebvre, Andrés Finzi, Madeleine Durand

**Affiliations:** University of Ottawa Heart Institute, Ottawa, Canada; Centre de recherche du CHUM, Montreal, Canada; Department of Radiology, Radiation Oncology and Nuclear Medicine, Université de Montréal, Montréal, Canada; Département of Medicine, Université de Montréal, Montreal, Canada; Département de microbiologie, infectiologie et immunologie, Université de Montréal, Canada

**Keywords:** HIV-1, soluble gp120, inflammation, pericoronary fat attenuation index, CT-scan, cardiovascular disease, Canadian HIV, Aging Cohort Study

## Abstract

Persistence of HIV antigens may drive chronic inflammation, leading to early-onset comorbidity among people living with HIV. We found that the presence of soluble glycoprotein 120 in plasma is associated with increased coronary inflammation, as measured by the pericoronary fat attenuation index (PFAI), a predictor of overt cardiovascular disease.

---

People living with HIV (PLWH) face an increased risk of cardiovascular disease (CVD) associated with *inflammaging*. ^1-3^ Despite suppressive antiretroviral therapy, residual production of HIV transcripts and proteins from the viral reservoir persists, driving chronic inflammation and immune activation.^4,5^ In that context, the HIV-1 envelop (Env) glycoprotein, particularly its gp120 subdomain, has been shown to exert diverse immunoregulatory effect. Using samples from participants from the Canadian HIV and Aging Cohort Study, we showed that ∼28% of PLWH virally suppressed on ART have measurable levels of soluble gp120 (sgp120), which are associated with increased inflammation, immune dysfunction and subclinical cardiovascular disease (CVD).^6^

Pericoronary Fat attenuation Index (PFAI) is a non-invasive imaging biomarker, which quantifies the level of inflammation present in peri-coronary adipose tissue, using its attenuation on computed tomography (CT).^7^ Lower attenuation (closer to -190 Hounsfield units [HU]) is associated with less inflammation, while higher values (closer to -30 HU) indicates more inflammation.^8^ PFAI has been shown to predict incident myocardial infarction, even after controlling for all traditional cardiovascular risk factors and coronary artery plaque caracteristics.^9^ We hypothesized that PFAI would be ideally suited to capture the inflammation associated with sustained sgp120 stimulation.

## METHODS

We performed a secondary analysis of the cardiovascular imaging sub-study of the CHACS, which has been published previously.^10,11^ We included participants living with HIV aged 40 years or more, with a baseline 10-year Framingham risk score ranging from 5 to 20% risk of cardiovascular disease at 10 years, an undetectable viral load, with available measurements for PFAI and sgp120. Covariates included socio-demographic and socioeconomic characteristics, traditional cardiovascular risk factors, and HIV-specific variables. PFAI was obtained using Siemens syngo.via Frontier software, by calculating the mean attenuation of all fat voxels (defined as those with an attenuation between - 190 and -30 HU) located within a radial distance from the outer vessel wall equal to the vessel’s diameter, in the segment of the right coronary artery situated 10 to 50 mm from the ostium, as recommended.^7^

Plasmatic sgp120 levels for each participant were previously measured. ^6^

Uni and multivariable linear regression models were then constructed to evaluate the crude and adjusted associations between presence of detectable sgp120 and PFAI. Statistical analyses were done using STATA version 17. All participants gave written informed consent, and the study was approved by the Research ethics board of the *Centre hospitalier de l’université de Montréal*. (#MP-02-2012-3068).

## RESULTS

The study included 145 PLWH (11 women and 134 men), with a mean age of 56.6 ± 6.5 years. White participants accounted for 84.8%, black participants for 8.2%. A total of 70.6 % of participants reported current or past tobacco use, while alcohol abuse was reported in 7.6 %. 31.5% of participants were hypertensive, 9% diabetic, and 21% on statin therapy. The mean Framingham risk score was 10% (xx to xx %). Participants had been living with HIV for a mean duration of 18.3 years, and were taking antiretroviral treatment for 14 years on average, and all had undetectable HIV viral load. Mean CD4 levels were 620 cells/mL, and the mean CD4/CD8 ratio was 0.96. The median CD4 nadir was 200, and 52.8% of participants reported a CD4+ nadir of less than 200 cells/ml.

Sgp120 was detectable in 65 participants (44.8%). The mean PFAI was -77.07 95%CI [-79.21 to - 74.93] in participants with detectable sgp120, and -80.35 95%CI [-82.55 to -78.15] in those without (p-value 0.037). Presence of CVD on CCTA was also associated with higher PFAI (-77 HU vs -81 HU, p=0.028). The figure shows the localization of pericoronary fat and PFAI measurements.

**Figure.**
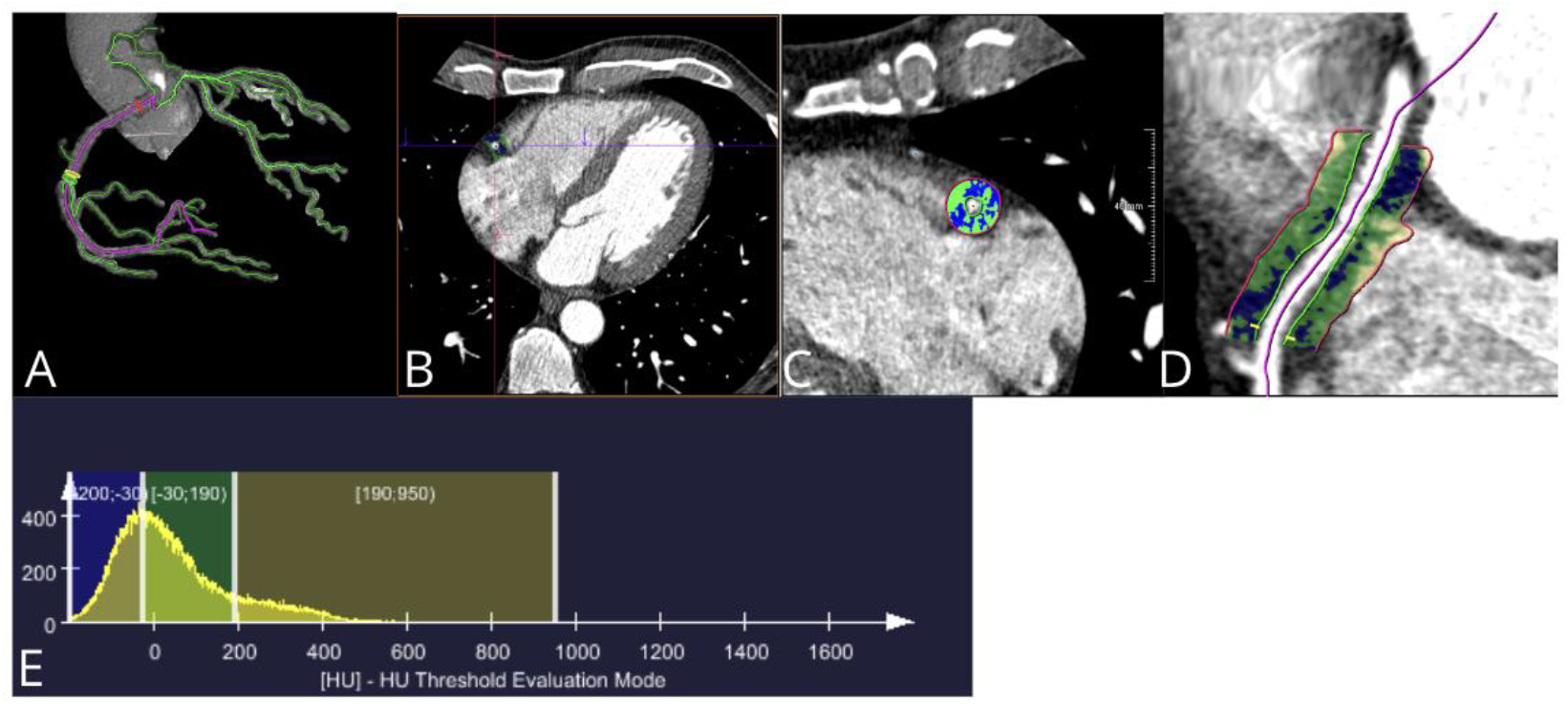
Post-processing of cardiac computed tomography angiography images to obtain PFAI A) Reconstruction of coronary arteries B) Right coronary artery C) cross-sectional region of interest of right coronary artery PFAI D) longitudinal region of interest for right coronary artery PFAI D) histogram of all fat voxels within region of interest

The results of uni and multivariable association between traditional, HIV related risk factors and PFAI are shown in the Table 1. Briefly, in the univariable models, female sex and statin treatment were associated with lower PFAI, suggesting lower cardiovascular disease risk, while current smoking and detection of sgp120 were associated to higher PFAI, indicating increased pericoronary fat inflammation. In the multivariable models, detectable sgp120 levels remained associated with increased PFAI, even after adjusting for statin use and smoking.

**Table.**
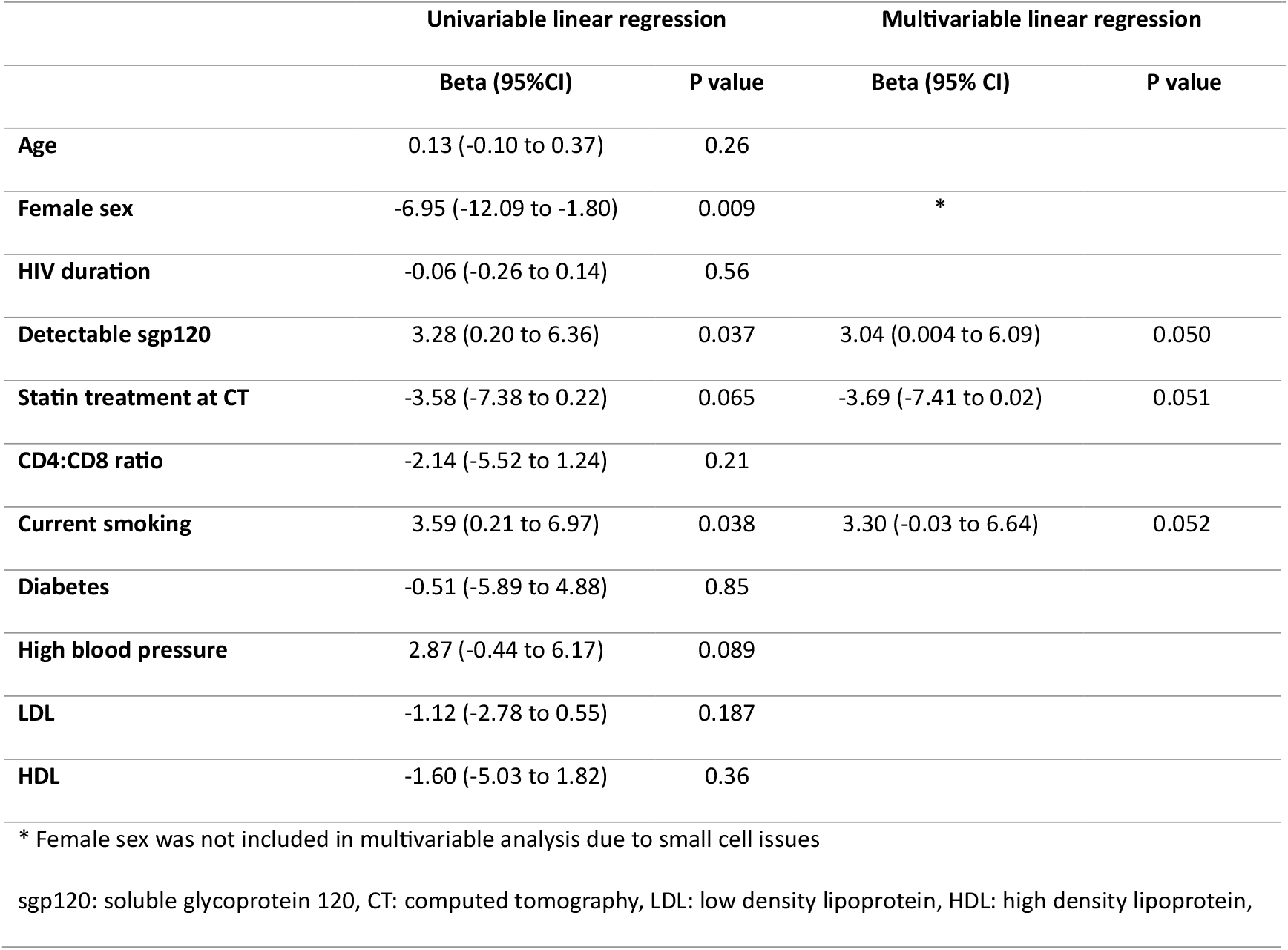
Crude and adjusted associations between risk factors and pericoronary fat attenuation index.

## DISCUSSION

In this cross-sectional analysis of the cardiovascular imaging sub-study of the Canadian HIV and Aging Cohort Study, we found that the persistence of sgp120 in plasma of PLWH with undetectable viral load was associated with increased inflammation in the pericoronary adipose tissue, as measured with PFAI. Interestingly, this association remained even after adjusting for statin treatment, suggesting that the chronic inflammation driven by viral persistence is not reversed with statin treatment.

Here we report the association between the HIV proinflammatory protein sgp120 and a clinically-validated measure of pericoronary adipose tissue inflammation, PFAI.

PFAI has emerged as a promising surrogate endpoint for CVD in PWH^12^, as it overcomes several limitations of coronary artery plaque analysis. It can detect disease across all CVD stages (even before the appearance of coronary artery plaque), and it reflects current inflammatory activity rather than long-term accumulated damage, making it more responsive to reversible risk factors like immune status and inflammation. PFAI also has strong predictive value for clinical events such as myocardial infarction even after accounting for traditional risks, and was recently found to predict all-cause mortality in PLWH.^12^ Statistically, PFAI presents normally-distributed continuous values, which has analytical advantages and offer optimal power.^13-15^ In keeping with this, PFAI has been shown to be sensitive to reductions in systemic inflammation associated with biologic treatment in a cohort of people with psoriasis arthritis.^16^ Combined, this suggests that PFAI may be used as a surrogate biomarker for cardiovascular risk reduction to evaluate novel therapeutic strategies for PLWH.

Our study has several limitations: a lack of women in the study sample, and its cross-sectional nature. A longitudinal study following the changes in PFAI and levels of sgp120 is warranted to underpin the exact mechanism of this interaction.

Currently, strategies to reduce CVD risk in PLWH are limited. The REPRIEVE trial showed that statins (pitavastatin) can lower the risk, but only in low-risk individuals, and without addressing the underlying virus-specific chronic inflammation present in PLWH.^17^ No treatment strategy currently targets the residual production of HIV antigens. Despite its limitations, our study supports the hypothesis that sgp120, even under suppressive ART, may contribute to cardiovascular disease in PLWH, and that it could represent a new therapeutic target for reducing the burden of ischaemic heart disease in PLWH.

In conclusion, our study supports the hypothesis that persistent HIV antigens contribute to increased cardiovascular risk for PLWH, even under suppressive ART, and supports future interventional studies targeting viral persistence to achieve successful aging with HIV.

## I. ACKNOWLEDGEMENTS

## II. AUTHORS’ CONTRIBUTUION

All authors contributed significant intellectual content to the study. PWHBT, BV and MD wrote the initial draft of the manuscript. KB, VS, BM and VDD performed PFAI measurements. MB performed sgp120 measurements. MB, JR and AF designed the sgp120 assay. CCL supervised cardiovascular imaging. MD did the statistical analysis. All authors revised the manuscript for critical intellectual content.

## III. DATA AVAILABILITY

Data is available upon reasonable request, in compliance with the data sharing procedures of the Canadian HIV and Aging Cohort Study.

